# Visual Outcome of Anti-Vascular Endothelial Growth Factor Injection at the University of Gondar Hospital Tertiary Eye Care and Training Centre, North-West Ethiopia

**DOI:** 10.1101/2022.08.23.22279127

**Authors:** Kassahun Endale, Asamere Tsegaw

## Abstract

**Background:** The management of macular edema and ocular neovascularization is changing and includes a new group of drugs called anti-vascular endothelial growth factor (anti-VEGF). Intra-vitreal injection of anti-VEGF agents has become the new standard of care for macular edema. However, data on their real world effectiveness and safety of these drugs in African eye care settings are very scarce.

**Objective:** To assess the visual outcome of intravitreal Avastin (IVA) injection at University of Gondar hospital tertiary eye-care and training center.

**Methods:** A retrospective analysis of medical records of patients who received IVA at the center was done. The main outcome measure was visual acuity (VA).

**Results:** The study included 37 eyes of 34 study participants with macular edema secondary to diabetic retinopathy, retinal vein occlusions, and neovascular age related macular degeneration (AMD). Mean VA improved from 6/60 (approximate 35 ETDRS letters) at baseline to 6/24 (approximate 55 ETDRS letters) at 2 months follow-up (*p=0*.*0045*) and this improvement was maintained at 6 months of follow up. This happened after mean injection of 2.5 times per eye over 6 months period. No major ocular or systemic treatment related adverse events were observed.

**Conclusion:** Patients who received IVA as initial therapy for macular edema from diabetic retinopathy, retinal vein occlusions, and neovascular AMD has a significant mean VA improvement which was maintained up to 6 months. Short term results show that IVA is effective and safe.

## Introduction

Macular edema and ocular neovascularization due to Retinal vascular diseases such as diabetic retinopathy and retinal vascular occlusions are major and growing causes of vision loss and blindness worldwide. Diabetic retinopathy remains the main cause of visual loss in the working age group in the developed world and is increasing as a major cause of blindness in developing countries (1, 2).

Retinal vascular occlusion, which includes Central retinal vein occlusion (CRVO) and branch retinal vein occlusion(BRVO), is the second most common cause of blindness from retinal vascular disease, second only to diabetic retinopathy (3, 4).

Vascular endothelial growth factor (VEGF) is the key molecular mediator of macular edema and ocular neovascularization due to retinal vascular diseases. VEGF inhibitors are now the first-line treatment offered to patients who have macular edema and intraocular neovascularization (3).

The inferior long-term results of alternative therapies, combined with an excellent safety profile from anti-VEGF treatment, make anti VEGF agents the current recommended first-line therapy for choroidal neovascularization (5).

In patients with macular edema secondary to retinal vein occlusive diseases, gender, patient’s age, systemic hypertension, baseline Best corrected visual acuity (BCVA), duration of veinous occlusion, perfusion status of the macula, and the number of injections were found to be of prognostic relevance for visual improvement at 6 months (6).

Avastin is a full-length humanized monoclonal antibody that binds to all isoforms of VEGF (5). Several studies done mainly in the western eye care settings have confirmed that intravitreal injection of Avastin is effective for the treatment of macular edema due to diabetic retinopathy and retinal vein occlusion and also for choroidal neovascularization due to Age Related Macular Degeneration (AMD). These studies have also confirmed intravitreal injection of Avastin has similar effectiveness and less expensive compared with newer anti-VEGF drugs like Ranibizumab and Aflibercept (1, 3, 4, 7, 8, 9, 10).

However, Avastin is still being used as an off-label drug and evidences on its effectiveness and safety in African eye care settings are scarce. This study attempts to assess the effectiveness and safety of intravitreal Avastin injection for ME due to retinal vascular diseases at University of Gondar tertiary eye care and training center.

## Materials and Methods

### Study design and period

A hospital based retrospective cross sectional study was done on patients who received IVA from August 2021-November 2021.

### Study area

This study was conducted at University of Gondar tertiary eye care and training center which is a major ophthalmic center in Ethiopia. It is an ophthalmic referral center for the entire North-West Ethiopia of an estimated 14 million people. Over 50,000 patients are seen at the center annually as inpatient and outpatient basis. Currently there are 6 subspecialty clinics with 7 actively working ophthalmologists, 26 ophthalmology trainee residents, 38 optometrists, 35 general clinical nurses and ophthalmic nurses and other supporting staff working in the center.

### Study population

All patients attending UOG hospital tertiary eye care and training center retina subspecialty clinic who have received intravitreal avastin injection in the study period and fulfilled the inclusion criteria.

### Inclusion and exclusion criteria

#### Inclusion criteria

All patients who received intravitreal avastin at UoG comprehensive specialized referral hospital, tertiary eye care and training center during the study period and had at least 2 months of follow up visits.

#### Exclusion criteria

Patients on follow after intravitreal Avastin injection elsewhere.

Patients who have intravitreal avastin injection but have a duration of follow up less than 2 months

Patients who have intravitreal steroid injection

Patients who have laser treatment

Patients with corneal and lenticular opacity

#### Operational definitions

**Vision Improved** – at least increments of VA by one line in the ETDRS chart

**Vision not improved -** at least decrement of VA by one line in the ETDRS chart or no change at all

#### Data collection tools and procedures

The surgical log book and patients’ medical chart was reviewed by using a structured checklist. Relevant information including Socio-demographic data, duration of visual complaint, which eye was involved, duration of follow up since the first injection, indication for injection, the dose of injection and the number of injections were reviewed and registered on the checklist. Baseline and post injection best corrected Visual acuity (VA) records, relevant anterior segment examination findings that were done using Zeiss slit lamp biomicroscope, and detailed stereoscopic examination findings of the posterior segment with 90D volk in a dilated pupil by a retina specialist were reviewed. OCT and fluorescein angiography were not done due to absence of these equipments at the center. VA records with Snellen fraction chart were converted to approximate ETDRS letters for analysis using a formula **approximate ETDRS= 85+50×log(Snellen fraction)**. The patients’ medical record charts and surgical reports were collected and marked to avoid duplication. The principal investigator checked the data collection and the completeness of the filled questionnaire and data extraction check list.

#### Data processing and analysis

Data completeness was checked on each data collection day by the principal investigator. Data clearance and cleaning was done before data entry to the computer. Data was entered to Epi-data 4.6 then exported to SPSS 25 statistical software where analysis was also performed. Descriptive findings were presented in terms of numbers, percentages, means, and standard deviation and displayed on tables, bar graphs, and pie charts. Paired-samples *t-test* analysis was performed and P-value of less than 0.05 was considered as statistically significant.

#### Ethical considerations

Ethical clearance was obtained from UOG ethical review board and permission to undergo this study was obtained from the department of ophthalmology. Informed consent was obtained from each patient and it was also made clear that the identity of patients and their records were kept confidential.

## Results

A total of 37 eyes of 34 study participants (24 right eyes & 13 left eyes) were included in this study. The mean age of the study participants was 50.68 +/- 11.2 years and range between 22 & 72 years. Fifty percent of the study participants were in the age group between 51-72 years. The majority of the study participants were male (n=19, 55.9%).

The right eye was most often injected among the study participants, 64.8% (n=24).Fifty one point four percent of the eyes(n=19) had baseline VA of 6/18-6/60 *(approximate ETDRS 61-35 letters)*.**(Figure-1**).

**Figure 1:**
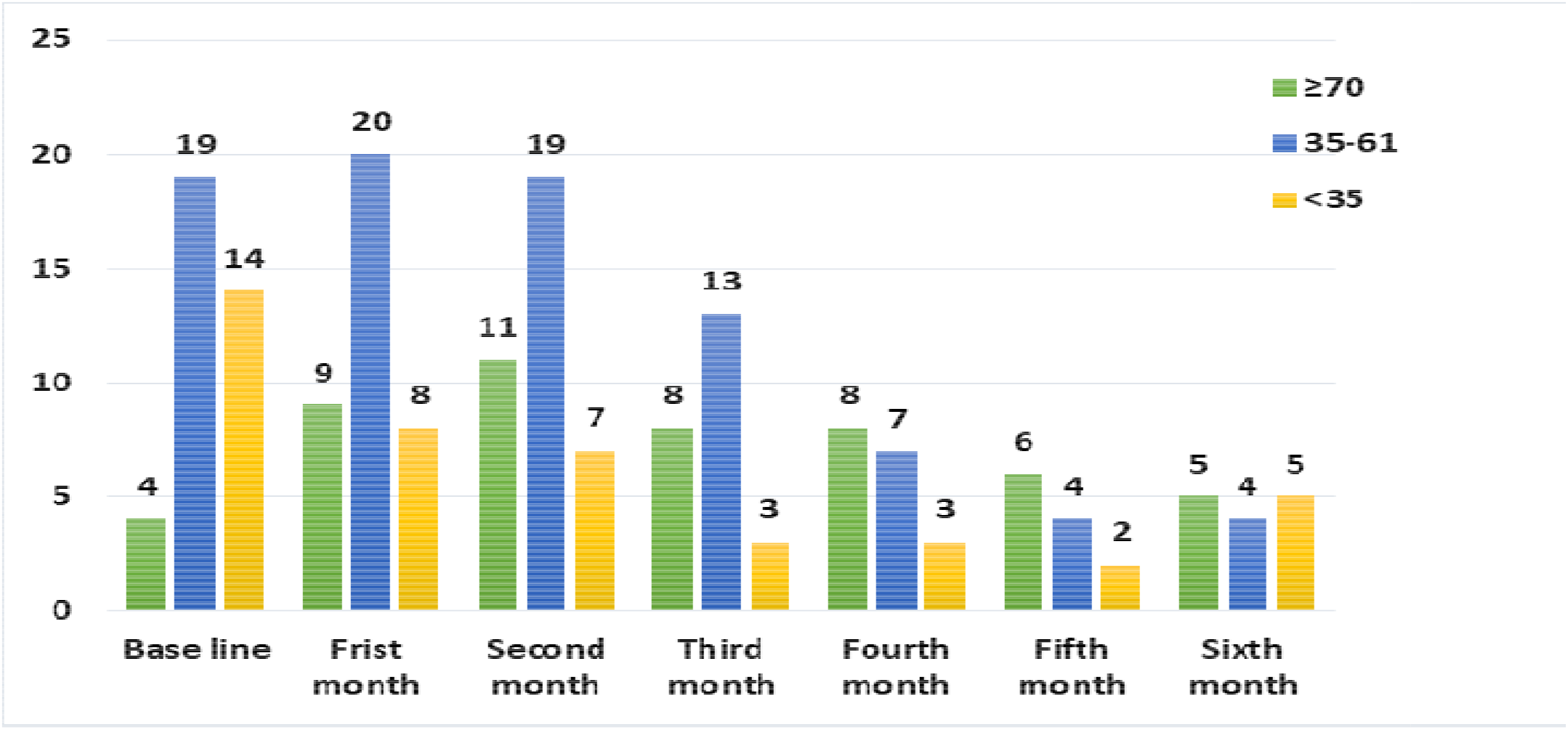
Baseline and Post IVA injection VA changes among study eyes at UoG tertiary eye care and training center retina sub-specialty clinic, Gondar, Ethiopia,2021.

The mean baseline VA among the study eyes was 35 ETDRS letters. Seventy five point seven percent(n=28) of the eyes received IVA after a mean duration of visual reduction of 16.36 weeks (range 1-52 weeks).Duration of visual reduction was not recorded in 24.3%(n=9) of the study eyes.**(Table-1)**

**Table 1:**
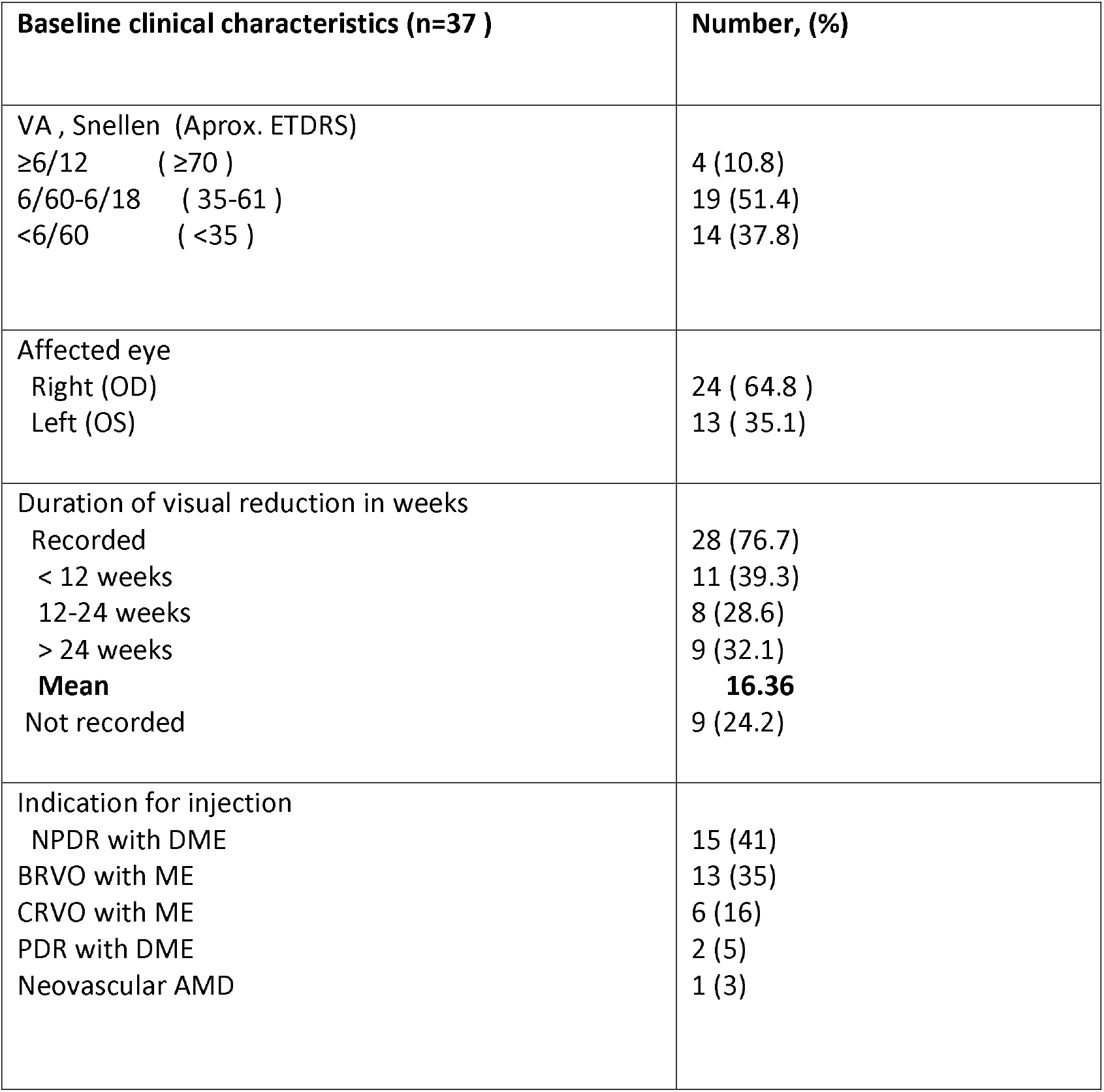
Baseline clinical characteristics of study eyes that were candidates for IVA injection at UoG tertiary eye care and training center retina sub-specialty clinic, Gondar, Ethiopia, 2021.

| Baseline clinical characteristics (n=37 ) | Number, (%) |
| --- | --- |
| VA , Snellen (Aprox. ETDRS)<br>≥6/12 ( ≥70 )<br>6/60-6/18 ( 35-61 )<br><6/60 ( <35 ) | 4 (10.8)<br>19 (51.4)<br>14 (37.8) |
| Affected eye<br>Right (OD)<br>Left (OS) | 24 ( 64.8 )<br>13 ( 35.1) |
| Duration of visual reduction in weeks<br>Recorded<br>< 12 weeks<br>12-24 weeks<br>> 24 weeks<br>**Mean**<br>Not recorded | 28 (76.7)<br>11 (39.3)<br>8 (28.6)<br>9 (32.1)<br>**16.36**<br>9 (24.2) |
| Indication for injection<br>NPDR with DME<br>BRVO with ME<br>CRVO with ME<br>PDR with DME<br>Neovascular AMD | 15 (41)<br>13 (35)<br>6 (16)<br>2 (5)<br>1 (3) |

One month after IVA injection, 59.3%(n=22) of eyes show ≥ 1 ETDRS line (5 ETDRS letters) improvement while at 2 months follow-up, 73%(n=27) of eyes show ≥ 1 ETDRS line (5 ETDRS letters)improvement from baseline. **(Figure-2)**

**Figure 2:**
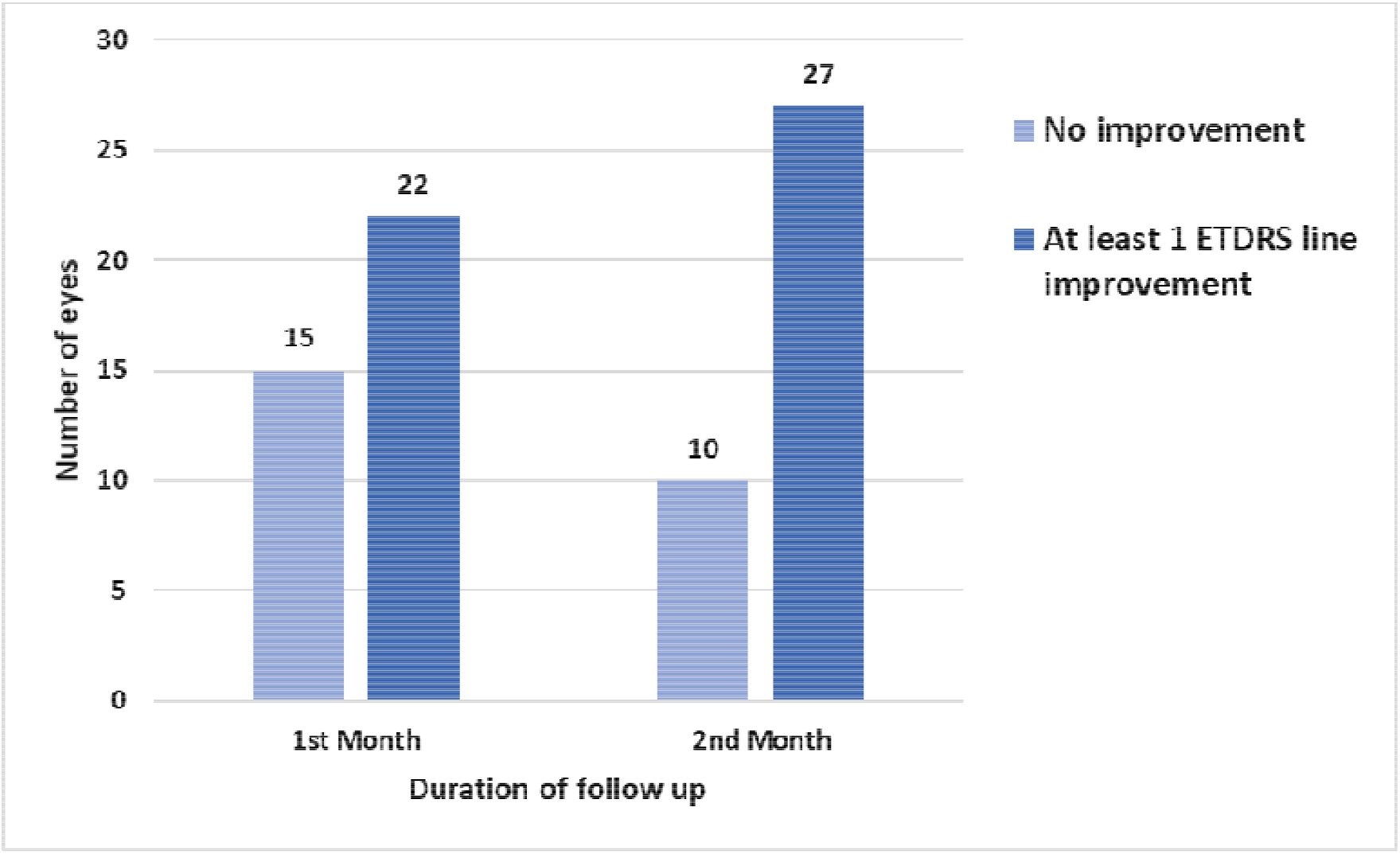
Bar graph showing post IVA injection response during the first 2 months among study eyes at UoG tertiary eye care and training center retina sub-specialty clinic, Gondar, Ethiopia, 2021.

The mean approximate ETDRS VA score improved from 35 letters at baseline to 45 letters(2 line improvement) at 1 month(10.61 letters,95% CI 3.56-17.7,*P=0*.*022)* and to 55 letters(4 line improvement from baseline) at 2 months (20 letters,95% CI 7.06-22.82, *p=0*.*0045*) was seen. This mean V/A improvement at 2 months is maintained throughout the study period and no statistically significant V/A improvement was seen after 2 months.**(Figure-3)**

**Figure 3:**
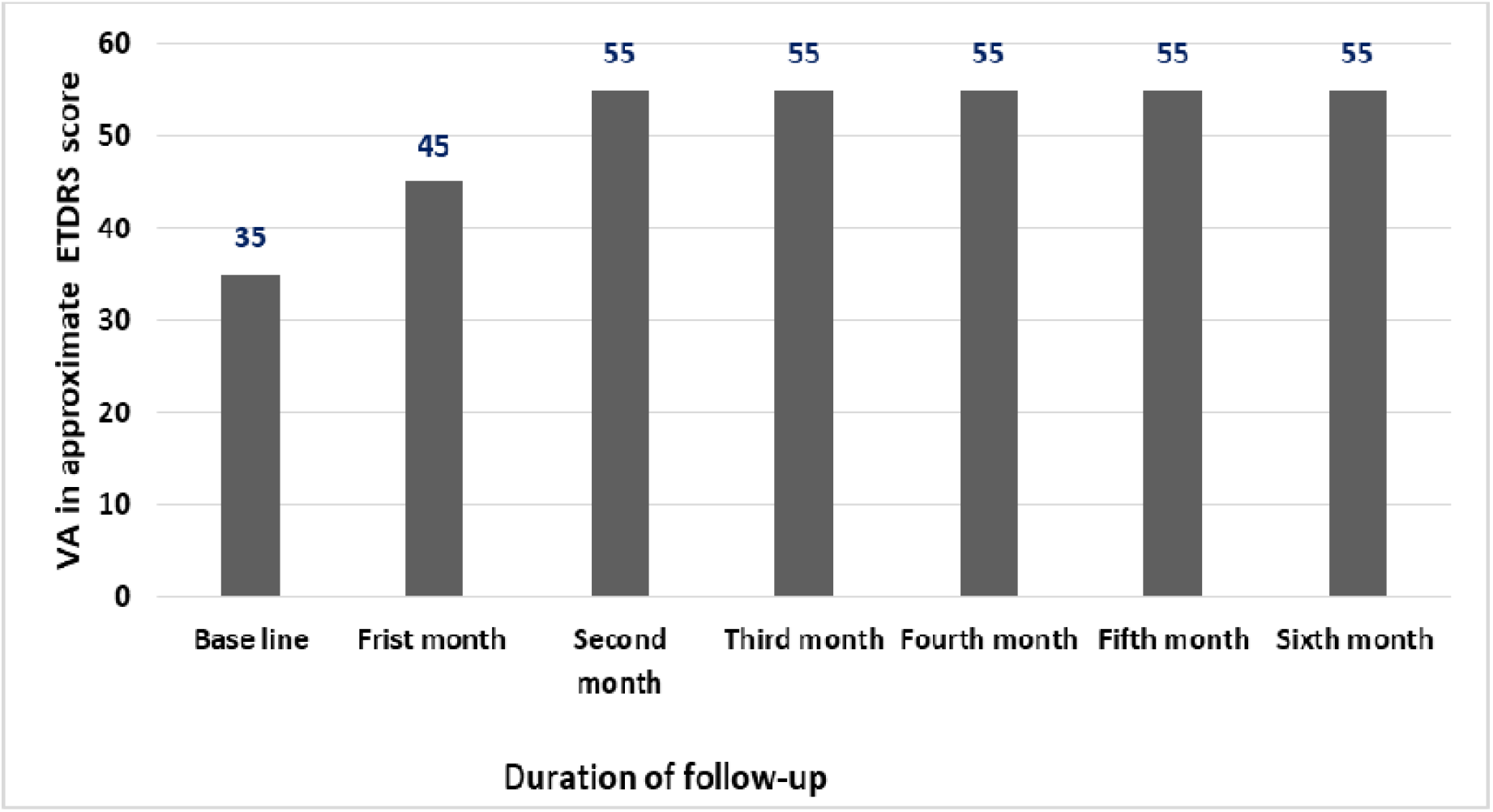
Baseline and Post IVA injection mean VA changes among study eyes at UoG tertiary eye care and training center retina sub-speciality clinic, Gondar, Ethiopia,2021.

### Sub-group VA pattern based on indication of IVA injection

Eyes with PDR with ME showed 1 ETDRS line decline from the baseline at the second month. Eyes with ME secondary to BRVO showed >4 ETDRS line improvement earlier than other eyes (in the second month) and at the 6^th^ month, there were 4 eyes with mean baseline and 6^th^ month vision of 26 & 35 respectively (2 ETRDS line improvement). Eyes with CRVO with ME have the worst baseline vision. (**Table-2)**

**Table-2:** Sub-group baseline and post IVA injection VA change among study eyes at UoG tertiary eye care and training center retina sub-specialty clinic, Gondar, Ethiopia,2021.

| Diagnosis | VA in approximate ETDRS score |  |  |  |  |  |  |
| --- | --- | --- | --- | --- | --- | --- | --- |
|  | Baseline mean ETDRS letters (n) | First month mean ETDRS letters (n) | Second month mean ETDRS letters (n) | Third month mean ETDRS letters (n) | Fourth month mean ETDRS letters (n) | Fifth month mean ETDRS letters (n) | Sixth month mean ETDRS letters (n) |
| NPDR with DME | 45 (15) | 55(15) | 55 (15) | 55(11) | 61 (7) | 61 (5) | 61 (6) |
| BRVO with ME | **30** (13) | **55**(13) | 61 (13) | 61(11) | 61(8) | 61(5) | **35** (4) |
| CRVO with ME | **15** (6) | 25 (6) | 25 (6) | 10 (2) | 25(3) | 5 (2) | 35 (3) |
| PDR with DME | 45 (2) | 45 (2) | **40** (2) | -- | -- | -- | -- |
| Neovascular AMD | 45 (1) | 45 (1) | 45 (1) | -- | -- | -- | 55 (1) |

The mean follow-up period among study eyes was 3.8 (range 2-6) months. There were 12 patients (14 eyes) who had follow visit at the 6^th^ month (35.3% of patients, 37.9% of eyes).However, only 7 eyes (18.9%) of 5 patients (14.7%) had consecutive monthly follow-up throughout the study period. **(Figure-4)**

**Figure 4:**
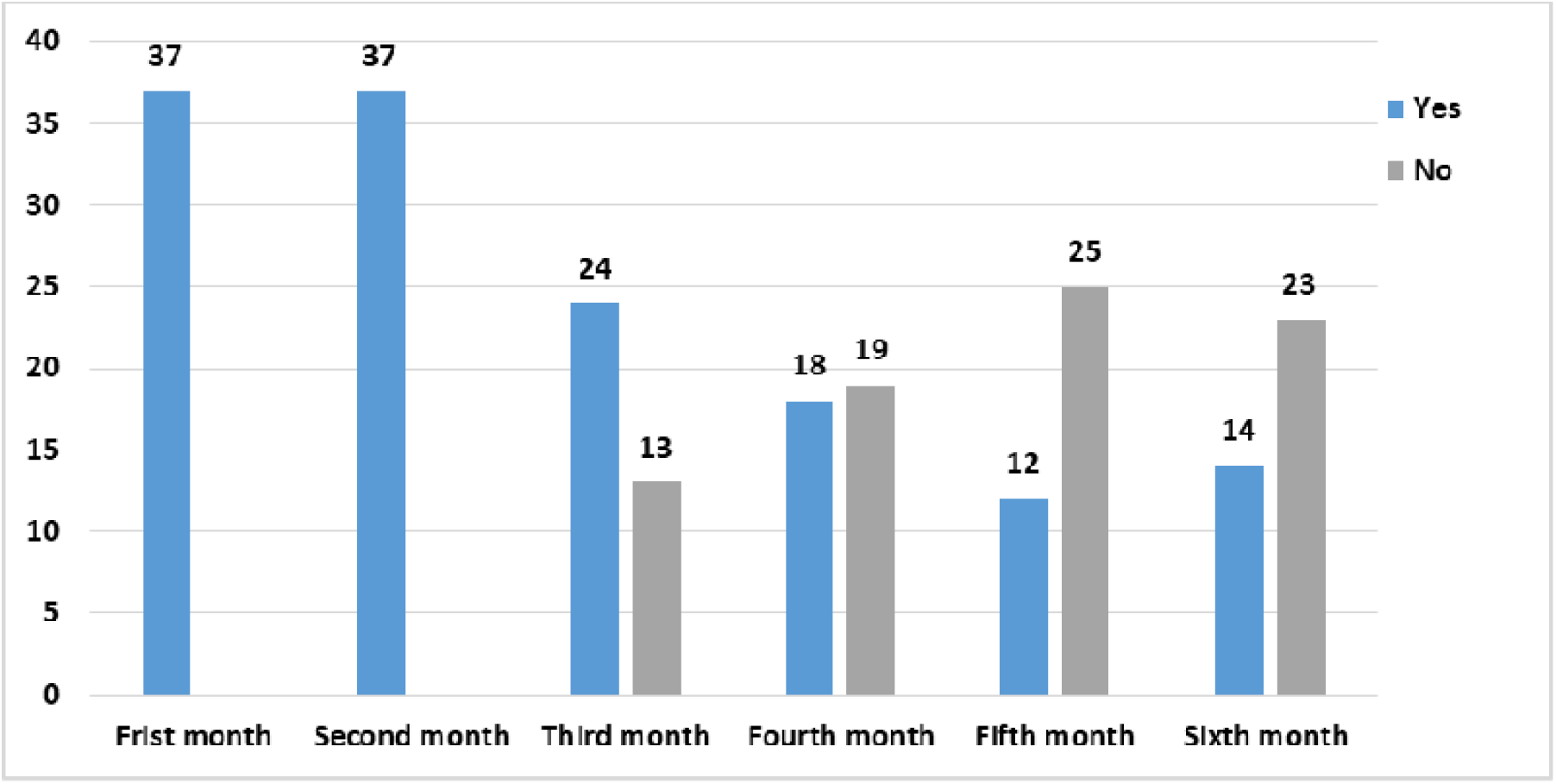
Post IVA injection follow-up visits among study eyes at UoG tertiary eye care and training center retina sub-specialty clinic, Gondar, Ethiopia,2021.

The total number of IVA injection among the study eyes was 92 (mean 2.5 injections per eye, range1-5).The majority of the eyes received 2 injections (45.9 %,n=17) and only 2 eyes received 5 injections (5.4%).**(Figure-5)**

**Figure 5:**
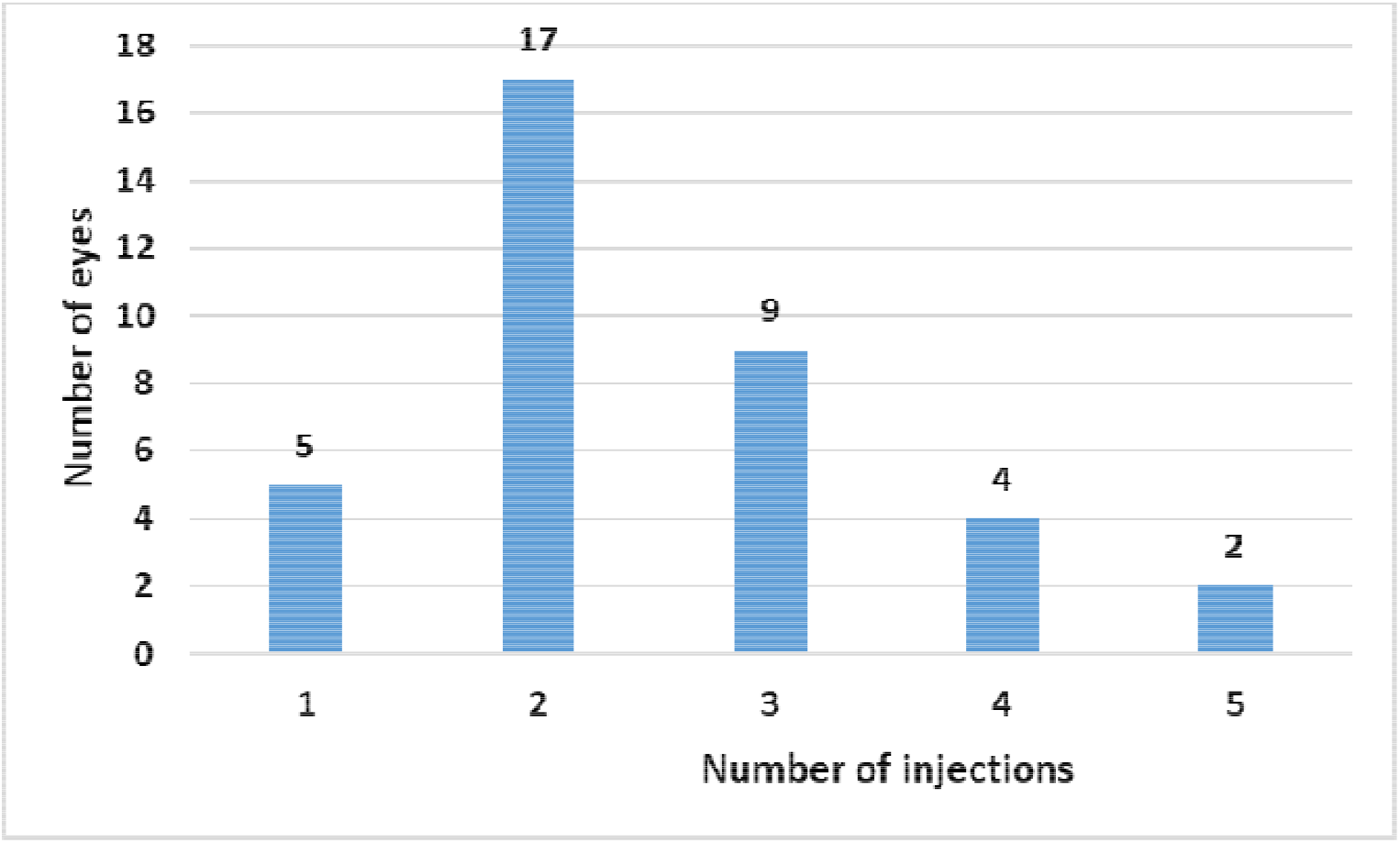
Bar graph showing the number of IVA injections among study eyes at UoG tertiary eye care and training center retina sub-specialty clinic, Gondar, Ethiopia,2021.

Common indications for IVA injection were macular edema secondary to NPDR(41%,n=15), BRVO (35%,n=13), CRVO (16%,n=6), PDR (5%,n=2), and neovascular AMD(3%,n=1).(**Figure-6)**

**Figure 6:**
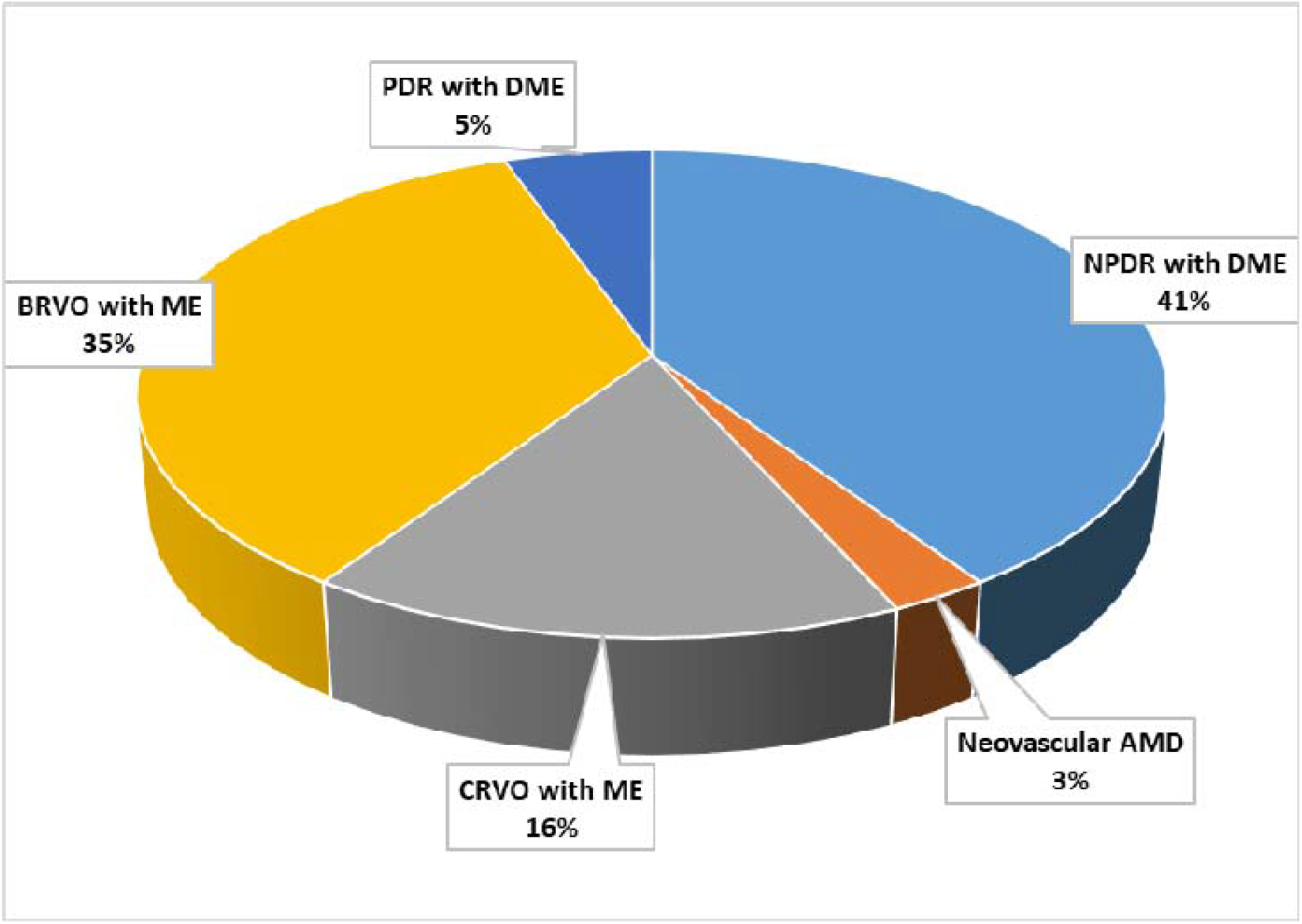
Common indications for IVA injection among study eyes at UoG tertiary eye care and training center retina sub-specialty clinic, Gondar, Ethiopia,2021.

## Discussion

The mean age of study participants was 50.68 ± 11.2 years and range between 22 & 72 years. This result is comparable to the result found in Korea where the mean age was 53.07±9.06 years among patients with BRVO with ME in whom comparative study of the visual outcome of the natural course group, IVA group, and intravitreal triamcinolone group was done. (11)

Studies done in patients who were about to undergo IVA injection for CRVO with ME and BRVO with ME in USA had older patients, 72.3 ±10 9 years, range 40-90 (3) and 65± 11.5 years respectively (12). This can be explained by the difference of life expectancy between ours and the American population.

Improvement in mean VA was seen after 1 month of IVA injection (approximately 10 ETDRS letters or 2 line improvement) from the baseline vision (*p=0*.*022*), which suggests that only a single injection of IVA may be effective at improving vision. However, it is unclear exactly how long the effect of just a single injection of IVA lasts and additional IVA injections are needed to maintain the improvement in VA. There is also significant Improvement of vision at 2 months of follow-up (20 ETDRS letters, or 4 line improvement from baseline, *p=0*.*0045*.This significant change of mean VA at 2 months follow-up is maintained throughout the 6-months follow-up. After 2 months follow-up, mean VA didn’t differ statistically. This is comparable with the result observed by the Pan-American collaborative Retina Study Group in 45 patients with ME secondary to BRVO who were treated, an average 2.5-injections per eye, and improved on average 4.8 ETDRS lines of VA from baseline at 6 months follow-up(12). This is also similar to the result found in Brazil, Mexico, Venezuela, and Colombia(1). A prospective study of 26 eyes of 26 patients with neovascular AMD in Denmark found that there was significant improvement in mean VA 6 weeks after IVA injection but no significant improvement in VA from baseline was found after 3 and 6 months (13).

The percentage of eyes that showed ≥ 1ETDRS line improvement at 1 month follow-up in this study (59.3%) from baseline compares favorably with the results of IVA injection in black patients with diabetic macular edema which showed a mean gain of 1ETDRS line improvement after a single injection only in 36% of eyes. This difference in response with ours may be due to the larger sample size in the mentioned study (14).

In this study, the most common indications for IVA injection were NPDR with DME and retinal vein occlusions with ME. Similar results were observed in studies done by Pan-American Collaborative Retina Study Group (1).

The mean number of IVA injection of our study participants was 2.5-injections per eye which is similar to that found by the Pan-American collaborative Retina Study Group (12) but fewer than other study results observed in USA where the mean number of injection for patients with ME secondary to CRVO was 5.9-injections per eye (3):and in Hong Kong where the mean number of injections was 3.53± 1.7 (range 3-10) for patients with choroidal neovascularization (15).This may be due to small sample size, shorter duration of follow-up, and loss to follow-up of our study participants.

The study reports only short-term VA changes after IVA injection relative to baseline vision but didn’t report anatomical changes or associated factors that determine treatment outcome.

The small sample size, non-protocol VA measurements performed using Snellen chart, lack of a control arm, and the retrospective design are some of the limitations of our study.

## Conclusion

The study result indicates that IVA is associated with mean VA improvement for patients with macular edema secondary to common retinal vascular diseases and neovascular AMD. A total of 92 IVA injections were given. None of the patients developed treatment-related vision threatening ocular complications such as endophthalmitis, retinal detachment, vitreous hemorrhage, cataract or systemic thrombo-embolic events.

## Data Availability

All relevant data are within the manuscript and its Supporting
Information files.

## Notes

### Competing Interest Statement

The authors have declared no competing interest.

### Funding Statement

no funding for this study

### Author Declarations

Ethical clearance was obtained from University of Gondar ethical review board and permission to undergo this study was obtained from the department of ophthalmology. Informed consent was obtained from each patient and it was also made clear that the identity of patients and their records were kept confidential.

## References

1. Arevalo JF, Fromow-Guerra J, Quiroz-Mercado H, Sanchez JG, Wu L, Maia M, et al. Primary intravitreal bevacizumab (Avastin) for diabetic macular edema: results from the Pan-American Collaborative Retina Study Group at 6-month follow-up. Ophthalmology. 2007;114(4):743–50.

2. Salam A, DaCosta J, Sivaprasad S. Anti-vascular endothelial growth factor agents for diabetic maculopathy. The British journal of ophthalmology. 2010;94(7):821–6.

3. Bajric J, Bakri SJ. Outcomes of Patients Initially Treated with Intravitreal Bevacizumab for Central Retinal Vein Occlusion: Long-Term Follow-Up. Seminars in ophthalmology. 2016;31(6):542–7.

4. Ehlers JP, Decroos FC, Fekrat S. Intravitreal bevacizumab for macular edema secondary to branch retinal vein occlusion. Retina (Philadelphia, Pa). 2011;31(9):1856–62.

5. Adatia FA, Luong M, Munro M, Tufail A. The other CNVM: a review of myopic choroidal neovascularization treatment in the age of anti-vascular endothelial growth factor agents. Survey of ophthalmology. 2015;60(3):204–15.

6. Jaissle GB, Szurman P, Feltgen N, Spitzer B, Pielen A, Rehak M, et al. Predictive factors for functional improvement after intravitreal bevacizumab therapy for macular edema due to branch retinal vein occlusion. Graefe’s archive for clinical and experimental ophthalmology = Albrecht von Graefes Archiv fur klinische und experimentelle Ophthalmologie. 2011;249(2):183–92.

7. van Asten F, Michels CTJ, Hoyng CB, van der Wilt GJ, Klevering BJ, Rovers MM, et al. The cost-effectiveness of bevacizumab, ranibizumab and aflibercept for the treatment of age-related macular degeneration-A cost-effectiveness analysis from a societal perspective. PloS one. 2018;13(5):e0197670.

8. Spooner K, Hong T, Fraser-Bell S, Chang AA. Current Outcomes of Anti-VEGF Therapy in the Treatment of Macular Oedema Secondary to Branch Retinal Vein Occlusions: A Meta-Analysis. Ophthalmologica. 2019;242(3):163–77.

9. Wu L, Arevalo JF, Roca JA, Maia M, Berrocal MH, Rodriguez FJ, et al. Comparison of two doses of intravitreal bevacizumab (Avastin) for treatment of macular edema secondary to branch retinal vein occlusion: results from the Pan-American Collaborative Retina Study Group at 6 months of follow-up. Retina (Philadelphia, Pa). 2008;28(2):212–9.

10. Emerson MV, Lauer AK, Flaxel CJ, Wilson DJ, Francis PJ, Stout JT, et al. Intravitreal bevacizumab (Avastin) treatment of neovascular age-related macular degeneration. Retina (Philadelphia, Pa). 2007;27(4):439–44.

11. Kim M, Jeong S, Sagong M. Efficacy of intravitreal bevacizumab for macular edema following branch retinal vein occlusion stratified by baseline visual acuity. Graefe’s archive for clinical and experimental ophthalmology = Albrecht von Graefes Archiv fur klinische und experimentelle Ophthalmologie. 2017;255(4):691–7.

12. Wu L, Arevalo JF, Berrocal MH, Maia M, Roca JA, Morales-Cantón V, et al. Comparison of two doses of intravitreal bevacizumab as primary treatment for macular edema secondary to central retinal vein occlusion: results of the pan American collaborative retina study group at 24 months. Retina (Philadelphia, Pa). 2010;30(7):1002–11.

13. Pedersen KB, Sjolie AK, Moller F. Intravitreal bevacizumab (Avastin) for neovascular age-related macular degeneration in treatment-naive patients. Acta ophthalmologica. 2009;87(7):714–9.

14. Osathanugrah P, Sanjiv N, Siegel NH, Ness S, Chen X, Subramanian ML. The Impact of Race on Short-term Treatment Response to Bevacizumab in Diabetic Macular Edema. American journal of ophthalmology. 2020;222:310–7.

15. Ng DS, Kwok AK, Tong JM, Chan CW, Li WW. Factors Influencing Need for Retreatment and Long term Visual outcome after Intravitreal Bevacizumab for Myopic Choroidal Neovascularization. Retina (Philadelphia, Pa). 2015;35(12):2457–68.

